# Distribution of residual disease in the peritoneum following neoadjuvant chemotherapy in advanced epithelial ovarian cancer and its potential therapeutic implications

**DOI:** 10.1101/2020.04.02.20048702

**Authors:** Aditi Bhatt, Naoual Bakrin, Praveen Kammar, Sanket Mehta, Snita Sinukumar, Loma Parikh, Sakina Shaikh, Suniti Mishra, Mita Y Shah, Vahan Kepenkian, Nazim Benzerdjeb, Olivier Glehen

## Abstract

**Introduction:** Residual disease in ‘normal appearing’ peritoneum is seen in nearly 30% following neoadjuvant chemotherapy for advanced epithelial ovarian cancer. Our goal was to study prospectively, the sequence of response in different regions, the commonest sites of occult residual disease, its incidence in different peritoneal regions and the potential therapeutic implications of these.

**Methods:** The patterns of response were studied based on the finding of residual disease in cytoreductive surgery specimens on pathological evaluation. A protocol for pathological evaluation was laid down and followed. Informed consent was taken from all patients. A correlation between clinical and pathological findings was made. Sugarbaker’s peritoneal cancer index was used to describe the regional distribution of peritoneal disease

**Results:** In 85 patients treated between July 2018 to June 2019, 83 FIGO stage III-C at diagnosis and 2 stage IV-A. Microscopic disease in ‘normal appearing’ peritoneal regions was seen in 22 (25.2%) and in normal peritoneum around tumor nodules in 30 (35.2%). Regions 4 and 8 of Sugarbaker’s peritoneal cancer index had the highest incidence of residual disease in absence of visible disease and regions 9 and 10 the lowest. The response to chemotherapy occurred in a similar manner in over 95% of the patients-the least common site of residual disease was the small bowel mesentery, followed by upper regions (regions 1-3), omentum and middle regions (regions 0, 4, 8), lower regions (regions 5-7) and lastly the ovaries. Nearly 85% had 4 or more peritonectomies and 67% had 6-7 peritonecomies.

**Conclusions:** Complete resection of involved the peritoneal region could address all the occult disease in a particular region. The role of resection of the entire region as well as ‘normal appearing’ parietal peritoneal regions (or total parietal peritonectomy) during interval cytoreduction should be prospectively evaluated to determine its impact on morbidity and survival.

## Introduction

Following a response neoadjuvant chemotherapy in advanced ovarian cancer, peritoneal deposits are replaced by areas of scarring or ‘normal appearing’ peritoneum that could be a potential site for residual disease in nearly 30% of the patients. [1] Though the goal of cytoreductive surgery in this setting is complete removal of all macroscopic disease, there is no clarity on the what to do with such areas. This is of importance as recurrence develops in nearly 75% of the patients in the peritoneum itself. [3, 4] In early stage disease, upstaging is seen in 18% as microscopic disease is seen in ‘normal appearing’ peritoneum when random biopsies are taken. [5, 6] This microscopic disease may respond to systemic chemotherapy. Following neoadjuvant chemotherapy, microscopic disease is that which has not been eradicated by chemotherapy and may contain chemotherapy resistant stem cells and may be a potential site of recurrence. [7] Secondly, when neoadjuvant chemotherapy is used for unresectable disease, there is peritoneal disease disease involving all parietal peritoneal surfaces. Thus, the parietal peritoneum has a greater potential to harbor residual disease since less than 10% have a complete response on pathology following neoadjuvant chemotherapy. [7] Current guidelines do not mention how much peritoneum to resect around tumor nodules. [8] They also recommend performing a biopsy/resection of suspicious areas. [9] Peritoneum involved prior to neoadjuvant chemotherapy in which disease has responded can be considered a ‘suspicious’ area and potential site of residual disease.

Following neoadjuvant chemotherapy, the ovaries and the omentum are the two commonest sites of residual disease. It is not known which are the other common sites of residual disease and if they should be resected when the visual appearance does not point toward disease.

Our goal was to prospectively study the pattern of response of peritoneal disease to neoadjuvant chemotherapy based on pathological evaluation of cytoreductive surgery specimens and the potential therapeutic implications. The sequence of response in different regions, most common sites of residual disease and the incidence of occult residual disease in different peritoneal regions was studied.

## Methods

This is a prospective study carried out at three peritoneal malignancy centers. The pathological findings in specimens of cytoreductive surgery were used to study the patterns of peritoneal dissemination. A protocol for pathological evaluation of these specimens was laid down and followed. Institutional and ethical approval was obtained. Surgical treatment and chemotherapy were performed as per the existing practices at each institution. Patients with advanced epithelial ovarian cancer (FIGO stage IIIC and IVA) having surgery following neoadjuvant chemotherapy were included in the study. Only those patients with extensive disease in which complete cytoreduction was not possible upfront received neoadjuvant chemotherapy at all centers. Patients were evaluated for surgery after 3 cycles of neoadjuvant chemotherapy. More cycles were given if disease was still unresectable. Some patients had an incomplete surgical procedure before neoadjuvant chemotherapy by non-specialist surgeons (those who are not gynecologic oncologists) and subsequently had a definitive procedure and these patients were also included in the study (referred to as secondary or ‘second look’ cytoreductive surgery here).

### Surgical intervention

All surgical procedures were performed with the goal of obtaining a complete cytoreduction (no visible residual disease). A thorough exploration of all regions was performed and extent of disease documented according to Sugarbaker’s peritoneal cancer index. This was the surgical peritoneal cancer index. [10] The extent of peritoneal resection was divided into 7 peritonectomy regions that is anteroparietal, pelvic, right upper quadrant, left upper quadrant, lesser omentum, greater omentum and mesenteric peritoneum. At two centers, complete removal of the parietal peritoneum was performed as part of an ongoing study approved by the ethical committees of both hospitals. Informed consent is taken from all patients. At the third center, such a protocol was not followed. However, some regions like the falciform, lesser omentum and umbilical round ligament were resected in absence of visible disease at this center too. Hence, regions in which the surgeon gave a lesion score of ‘0’ were considered uninvolved or ‘normal appearing’. The size of residual disease was recorded according to the completeness of cytoreduction score (CC-score). [10] Lymphadenectomy was performed if the nodes were enlarged or suspicious on imaging or during surgery.

### Pathological evaluation

The protocol for pathological evaluation was for peritonectomy specimens for which currently no guidelines exist and is described elsewhere. [11] The primary ovarian tumour and lymph nodes were analysed and reported as per the existing guidelines. [12] Briefly, each area of the peritoneum was marked in the en-bloc specimen by the surgeon and sent to the pathologist or sent as a separate specimen. The size of the largest deposit in each region was recorded. One or more sections were taken from the largest nodule. One or more sections were taken from peritoneum around tumour nodules showing no gross disease depending on the size of the resected specimen. The distance was 1 cm or more from the tumor nodule.

Appropriate immunohistochemistry markers were used to confirm the presence of disease when required. The pathological peritoneal cancer index was calculated on the lines of surgical peritoneal cancer index. [13] Nodes dissected with the resected segments of bowel, omenta and retroperitoneal nodes when dissected were evaluated for presence of disease.

### Disease distribution in the peritoneal cavity

The PeRitOneal MalIgnancy Stage Evaluation online application (e-PROMISE) was used to define anatomical structures in each region of the peritoneal cancer index (Figure 1). [14] The peritoneal cavity was divided into 3 regions-the upper region comprising of regions 1, 2 and 3, middle region comprising of regions 0, 4, 8 and the lower region comprising of regions 5, 6 and 7. The omentum was evaluated separately from the structures in region 0 and each small bowel region (9-12) as well. Histological confirmation of disease was taken as presence of tumour in that region.

**Figure 1.**
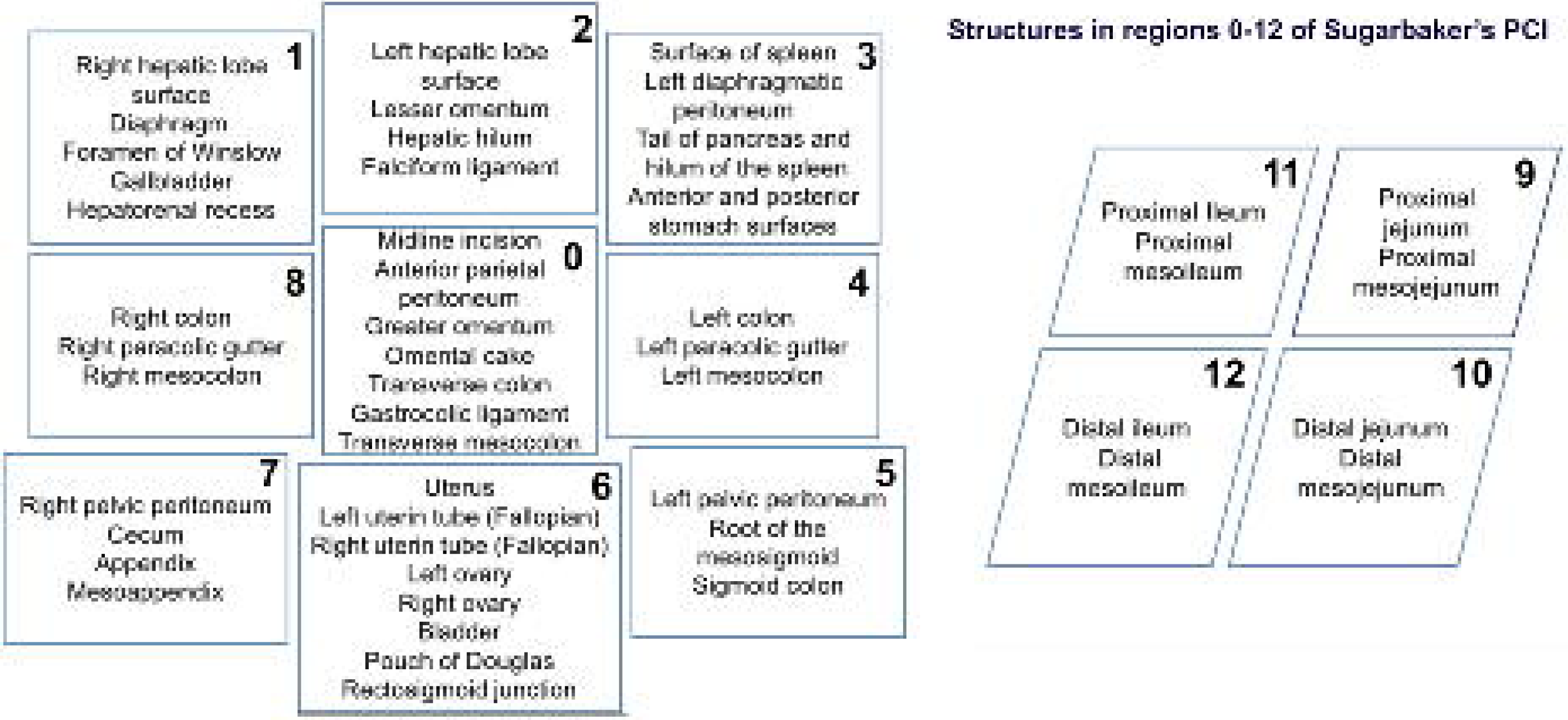
List of structures in each of the 13 regions of Sugarbaker’s PCI defined using the PROMISE internet application

### Pathological response to chemotherapy

The pathological response to chemotherapy was graded based on the chemotherapy response score developed by Bohm et al. **(Supplement 1)**. [15] The term ‘chemotherapy response grade’ is used here. The response was assessed not just in the ovaries and the omentum (as in Bohm’s study) but also at the peritoneal sites. [15]

### Statistical analysis

Categorical data were described as number (%). Abnormally distributed continuous data were expressed as the x^2^median and range. Categorical data were compared with the x^2^test. For comparison of median values, non-parametric independent sample t test and for means, independent sample t test was used, where in Levene’s test for equality of variance was applied. A p-value of <0.05 was considered statistically significant.

## Results

From July 2018 to June 2019, 85 patients undergoing interval or secondary CRS were included. Eighty-three patients had FIGO stage III-C at diagnosis and 2 had stage IV-A. Seventy-two (84.7%) patients underwent interval cytoreduction and 13 (15.3%) underwent secondary cytoreduction. The median surgical PCI was 12 [range-0-37]. A complete cytoreduction was obtained in 71 patients (83.5%) and and CC-1 resection in 12 (14.1%). Nearly 85% of the patients had 4 or more peritonectomies and 67% had 6-7 peritonecomies **(Table 1)**.

**Table 1.**
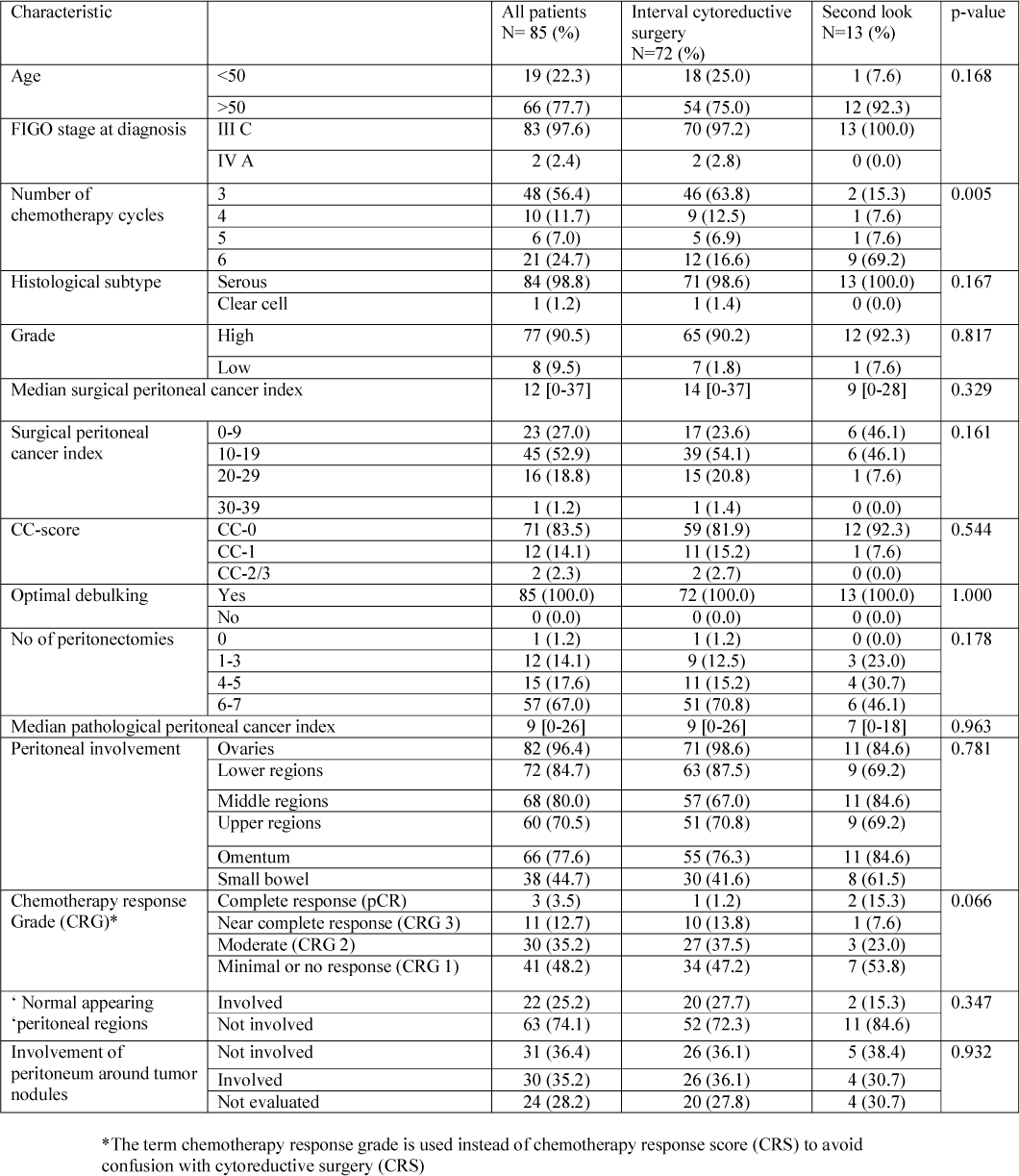
Clinical, surgical and pathological findings in 85 patients undergoing interval cytoreductive surgery

### Pathological findings

Three patients (3.5%) had a pathological complete response and 11(12.7%) had a near complete response **(Table 1)**. Microscopic disease in ‘normal appearing’ or uninvolved peritoneal regions (lesion score 0) was seen in 22 (25.2%). Microscopic disease in normal peritoneum around tumor nodules was seen in 30 (35.2%). The presence of microscopic disease in absence of visible disease in each region in shown in **Table 2**. Its incidence was the highest in regions 8 (11.2% of the patients in which the region was dissected) and 4 (10.0%) and the lowest in regions 9 (2.0%) and 10 (1.9%). Some had microscopic disease in absence of visible disease in more than one region. We compared patients with and without microscopic disease in ‘normal appearing’ areas and found no significant difference in the disease extent and distribution, surgical and pathological PCI, the number of peritonectomies and visceral resections performed **(Table 3)**. A larger proportion of patients who did not have disease in ‘normal appearing’ regions, received all 6 cycles of neoadjuvant chemotherapy (p=0.025)..

**Table 2.** Incidence of involvement of different peritoneal regions (according to Sugarbaker’s peritoneal cancer index) in absence of visible disease

| Peritoneal region | Dissection performed (%) | Not involved in absence of visible disease (%) | Involved in absence of visible disease |  | Total number in which it was involved (%) |
| --- | --- | --- | --- | --- | --- |
|  |  |  | In all patients (%) | In those which it was dissected (%) |  |
| 0 | 85 (100.0%) | 77 (90.5%) | 8 (9.5) | 8 (9.5) | 66 (77.6%) |
| 1 | 79 (92.9%) | 76 (89.4%) | 3 (3.5) | 3 (3.9) | 53 (62.3%) |
| 2 | 82 (96.4%) | 74 (87.0%) | 8 (9.5) | 8 (9.7) | 38 (44.7%) |
| 3 | 72 (84.7%) | 68 (80.0%) | 4 (4.7) | 4 (5.5) | 30 (35.2%) |
| 4 | 70 (82.3%) | 63 (74.1%) | 7 (8.2) | 7 (10.0) | 29 (34.1%) |
| 5 | 78 (91.7%) | 75 (88.2%) | 3 (3.5) | 3 (3.8) | 38 (44.7%) |
| 6 | 83 (97.6%) | 79 (92.9%) | 5 (5.8) | 5 (6.0) | 68 (80.0%) |
| 7 | 77 (90.5%) | 73 (85.8%) | 4 (4.7) | 4 (5.1) | 40 (47.0%) |
| 8 | 71 (83.5%) | 63 (74.1%) | 8 (9.5) | 8 (11.2) | 36 (42.3%) |
| 9 | 50 (58.8%) | 49 (57.6%) | 1 (1.2) | 1 (2.0) | 10 (11.7%) |
| 10 | 51 (60.0%) | 50 (58.8%) | 1 (1.2) | 1 (1.9) | 5 (5.8%) |
| 11 | 58 (68.2%) | 56 (65.8%) | 2 (2.3) | 2 (3.4) | 12 (14.1%) |
| 12 | 65 (76.4%) | 62 (72.9%) | 3 (3.5) | 3 (4.6) | 21 (24.7%) |

**Table 3.**
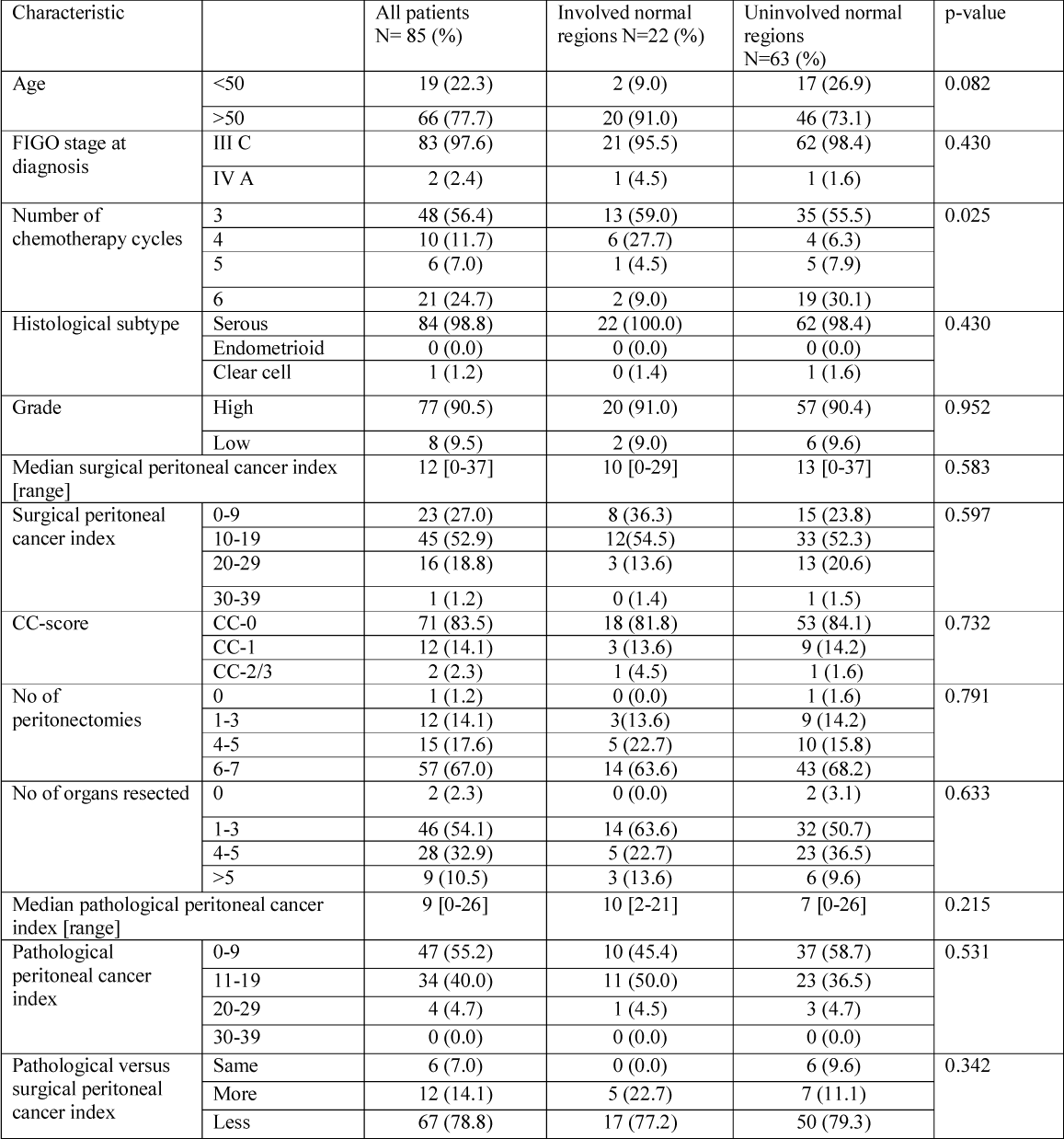
Comparison of clinical, surgical and pathological findings in patients with and without involvement of ‘normal looking’ peritoneal regions

| Characteristic |  | All patients<br>N= 85 (%) | Involved normal<br>regions N=22 (%) | Uninvolved normal<br>regions<br>N=63 (%) | p-value |
| --- | --- | --- | --- | --- | --- |
| Age | <50 | 19 (22.3) | 2 (9.0) | 17 (26.9) | 0.082 |
|  | >50 | 66 (77.7) | 20 (91.0) | 46 (73.1) |  |
| FIGO stage at<br>diagnosis | III C | 83 (97.6) | 21 (95.5) | 62 (98.4) | 0.430 |
|  | IV A | 2 (2.4) | 1 (4.5) | 1 (1.6) |  |
| Number of<br>chemotherapy cycles | 3 | 48 (56.4) | 13 (59.0) | 35 (55.5) | 0.025 |
|  | 4 | 10 (11.7) | 6 (27.7) | 4 (6.3) |  |
|  | 5 | 6 (7.0) | 1 (4.5) | 5 (7.9) |  |
|  | 6 | 21 (24.7) | 2 (9.0) | 19 (30.1) |  |
| Histological subtype | Serous | 84 (98.8) | 22 (100.0) | 62 (98.4) | 0.430 |
|  | Endometrioid | 0 (0.0) | 0 (0.0) | 0 (0.0) |  |
|  | Clear cell | 1 (1.2) | 0 (1.4) | 1 (1.6) |  |
| Grade | High | 77 (90.5) | 20 (91.0) | 57 (90.4) | 0.952 |
|  | Low | 8 (9.5) | 2 (9.0) | 6 (9.6) |  |
| Median surgical peritoneal cancer index<br>[range] |  | 12 [0-37] | 10 [0-29] | 13 [0-37] | 0.583 |
| Surgical peritoneal<br>cancer index | 0-9 | 23 (27.0) | 8 (36.3) | 15 (23.8) | 0.597 |
|  | 10-19 | 45 (52.9) | 12 (54.5) | 33 (52.3) |  |
|  | 20-29 | 16 (18.8) | 3 (13.6) | 13 (20.6) |  |
|  | 30-39 | 1 (1.2) | 0 (1.4) | 1 (1.5) |  |
| CC-score | CC-0 | 71 (83.5) | 18 (81.8) | 53 (84.1) | 0.732 |
|  | CC-1 | 12 (14.1) | 3 (13.6) | 9 (14.2) |  |
|  | CC-2/3 | 2 (2.3) | 1 (4.5) | 1 (1.6) |  |
| No of<br>peritonectomies | 0 | 1 (1.2) | 0 (0.0) | 1 (1.6) | 0.791 |
|  | 1-3 | 12 (14.1) | 3 (13.6) | 9 (14.2) |  |
|  | 4-5 | 15 (17.6) | 5 (22.7) | 10 (15.8) |  |
|  | 6-7 | 57 (67.0) | 14 (63.6) | 43 (68.2) |  |
| No of organs resected | 0 | 2 (2.3) | 0 (0.0) | 2 (3.1) | 0.633 |
|  | 1-3 | 46 (54.1) | 14 (63.6) | 32 (50.7) |  |
|  | 4-5 | 28 (32.9) | 5 (22.7) | 23 (36.5) |  |
|  | >5 | 9 (10.5) | 3 (13.6) | 6 (9.6) |  |
| Median pathological peritoneal cancer<br>index [range] |  | 9 [0-26] | 10 [2-21] | 7 [0-26] | 0.215 |
| Pathological<br>peritoneal cancer<br>index | 0-9 | 47 (55.2) | 10 (45.4) | 37 (58.7) | 0.531 |
|  | 11-19 | 34 (40.0) | 11 (50.0) | 23 (36.5) |  |
|  | 20-29 | 4 (4.7) | 1 (4.5) | 3 (4.7) |  |
|  | 30-39 | 0 (0.0) | 0 (0.0) | 0 (0.0) |  |
| Pathological versus<br>surgical peritoneal<br>cancer index | Same | 6 (7.0) | 0 (0.0) | 6 (9.6) | 0.342 |
|  | More | 12 (14.1) | 5 (22.7) | 7 (11.1) |  |
|  | Less | 67 (78.8) | 17 (77.2) | 50 (79.3) |  |

### Pattern of response

The ovaries (96.4%) were the most common site of residual disease followed by lower regions (84.7%), middle region (80.0%) and omentum (77.6%), upper regions (70.5%) and lastly small bowel sites (44.7%). The involvement of each small bowel region was low with region 9 involved in 11.7%, 10 in 5.8%, 11 in 14.1% and 12 in 24.7%. Small bowel involvement was not seen in absence of disease in both upper and lower regions. Residual disease in lower regions was not seen in 10 patients (11.7%). In most of these patients there was no residual disease in the upper and middle regions as well. Three patients (3.5%) had residual disease in the upper region in absence of disease in the lower regions, however, the ovaries had residual disease with surface involvement in all. Similarly, 5 patients (5.8%) had disease in the omentum (middle regions) in absence of disease in the lower regions but ovarian involvement was seen as described above. Thus, in nearly 95% of the patients, the omentum and upper regions were not involved in absence of pelvic peritoneal disease and in 100% they were not involved in absence of residual disease in the ovaries. Greater omental involvement was seen in 66 patients (77.6%) and in these the right upper quadrant was involved in 49 (74.2%). Disease in the upper region was not seen in absence of omental disease. In 53 (62.3%) patients with right upper quadrant (region 1) involvement, 28 patients (>50%) had region 3 involvement. Region 9 was involved in 4 (7.2%), 10 in 4 (7.2%), 11 in 10 (18.1%) and 12 in 14 (25.4%) of the 53 patients. Region 6 was involved in all these 53 patients.

### Lymph node involvement

Pelvic lymph nodes were dissected in 55 (74.8%) and involved in 22 (25.2%) **(Supplement 2)**. Para-aortic nodes were dissected in 41 (48.2%) and were involved in 6 (7.0%). Visceral nodes were dissected in 31 (36.4%) patients and positive in 6 (7.0%). None of the patients with a complete or near complete pathological response to chemotherapy had involvement of regional nodes.

## Discussion

This study brings to light the need to evaluate the role of performing wider resection of the involved peritoneal region in the setting of interval cytoreductive surgery. Microscopic disease in the normal peritoneum around tumor nodules was seen in 35.2% of the patients. Instead of wide resection of the involved peritoneum, resection of the entire peritoneal region may be able to address occult disease better and its impact on survival should be studied **(Figure 2)**. [16] Similarly, disease was seen in one or more uninvolved regions in 25.2% which suggests that the role of resecting uninvolved peritoneal regions should be evaluated prospectively.

**Figure 2.**
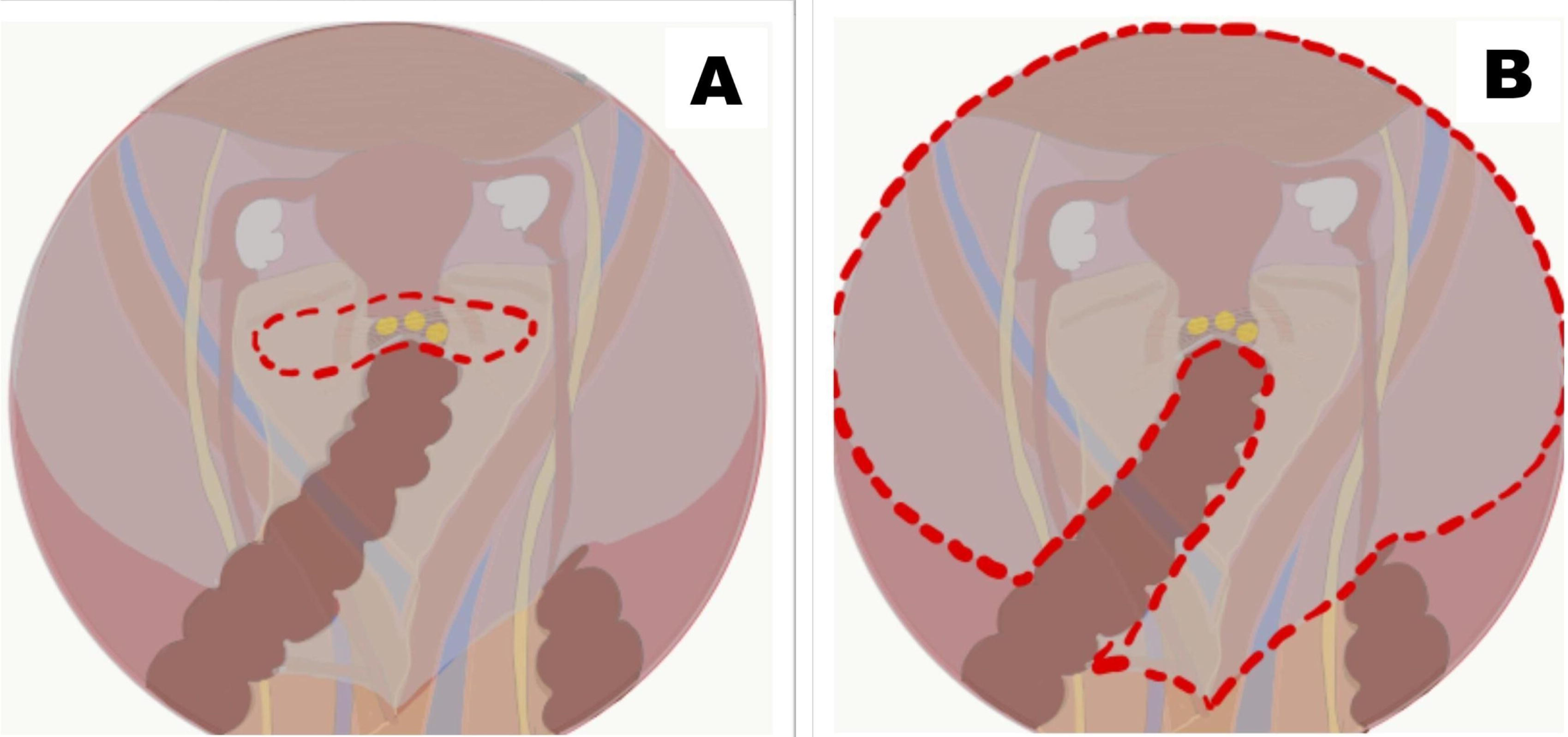
A: Wide resection of the tumour bearing peritoneum that is performed routinely; B: resection of the entire pelvic peritoneum to account for microscopic disease

### Resection of uninvolved peritoneal regions and pattern of response

There are some studies that have compared resection of only involved regions of the peritoneum (selective parietal peritonectomy) with resection of the entire parietal peritoneum (total parietal peritonectomy). [17, 18] Each of these has several limitations due to which nothing definite can be inferred from them. However, the morbidity of a total parietal peritonectomy was acceptable. Resection of uninvolved regions should be performed not just based on the probability of finding microscopic disease but also the the patterns of recurrence and its potential impact on survival.

It has been observed, the the parietal peritoneum is involved first and has a greater disease burden compared to the visceral peritoneum (excluding the omentum). One previous study showed that the visceral peritoneum responds first-the disease responds completely in the visceral regions (except the omentum) first followed by the parietal regions. [1] This study showed the same. The response to chemotherapy occurred showed a similar sequence in nearly 95%. The ovaries were the most common site of residual disease and the last to have a complete response, followed by lower regions, the omentum and middle regions, followed by the upper regions. The small bowel was the last site to have disease and the first to have a complete response. In this study, small bowel regions were dissected in 55-75% but involved only in 5 to 25%.

### Addressing residual disease after neoadjuvant chemotherapy

In ovarian cancer, a thorough exploration of the peritoneal cavity is needed to completely rule out sites of residual disease. Presence of gross/visible disease in one region could alert the surgeon of disease in another region. For e.g in patients with omental disease, pelvic regions were involved in 95% and the right upper quadrant in nearly 75% of the patients. Hence, these two regions should be dissected in such patients or at least thoroughly explored.

The use of neoadjuvant chemotherapy in epithelial ovarian cancer has been associated with an inferior survival. [19, 20, 21] One of the reasons for this could be that surgery targets only resection of sites of gross residual disease. It is most likely that a lot of microscopic disease is left behind when areas of scarring and/or previously involved areas are not resected and they are known to contain chemotherapy resistant cells. [7] When surgery is performed only targeting residual disease, survival outcome becomes largely dependent on tumor sensitivity to systemic chemotherapy. It is possible that more extensive surgery following neoadjuvant chemotherapy can in part offset the poor prognostic impact of a poor response to systemic chemotherapy and this should be further evaluated. [22]

### Addressing regional nodes

Though a recent randomized trial showed no benefit of systematic lymphadenectomy when nodes were not enlarged, this trial is only for surgery performed upfront and not for surgery following neoadjuvant chemotherapy. [23] Hence, in our opinion, there is no clear directive as to which patients should have regional lymphadenectomy following neoadjuvant chemotherapy. And residual disease in nodes after chemotherapy is not the same as in patients undergoing primary cytoreduction. In our study, pelvic nodes were involved in 25% and other nodes in 7-15%. After the peritoneum, regional nodes are the second most common site for disease recurrence/ progression. [24] And a reason for this could be the failure to address these nodes during surgery. The correlation between pelvic peritoneal disease burden and regional node involvement has been demonstrated. [25, 26, 27] Future studies should look at the correlation with peritoneal disease extent and impact of lymphadenectomy on survival in this setting.

Some of the strengths of this study are that majority of the patients had serous carcinoma with stage IIIC disease that was not amenable to complete macroscopic resection at initial presentation and the study was performed prospectively. It may be argued that different surgical protocols were used at different centers, but our goal was not to evaluate the role of less or more extensive surgery; it was to study the residual disease distribution. We enrolled consecutive patients being treated at different centers during a given time period for it. The other debatable point is describing our findings as ‘sequence of response’ since the evaluation is performed at one time-point only. The limitation of imaging in evaluating peritoneal disease extent has been demonstrated before. It would be ideal to have a laparoscopic evaluation of disease extent before each cycle of chemotherapy but that is not possible and in extensive disease, laparoscopy may not be able to inspect all areas. At the time of surgery, different patients are in different stages of response and since our findings show the same pattern in over 95% of the patients, it can be extrapolated that the response occurs in a sequential manner as we have described. Categorization of the morphology of the tumor deposits should be done in future studies.

## Conclusions

A significant proportion of patients undergoing interval cytoreductive surgery have microscopic residual disease in ‘normal appearing’ peritoneal areas and in the peritoneum surrounding tumor nodules. The parietal peritoneal regions are the more common sites of residual disease compared to visceral regions except the omentum and response to neoadjuvant chemotherapy occurs in a similar manner in most patients. These findings warrant a prospective evaluation of role of resection of the entire peritoneal region as well as resection of involved parietal regions (or total parietal peritonectomy) during interval cytoreduction. The role of resection of regional nodes in relation to the peritoneal disease should be evaluated prospectively.

## Data Availability

Data is available on reasonable request

## References

1. Bhatt A, Sinukumar S, Mehta S, et al. Patterns of pathological response to neoadjuvant chemotherapy and its clinical implications in patients undergoing interval cytoreductive surgery for advanced serous epithelial ovarian cancer-A study by the Indian Network for Development of Peritoneal Surface Oncology (INDEPSO). Eur J Surg Oncol. 2019 Apr;45(4):666–671. doi: 10.1016/j.ejso.2019.01.009. Epub 2019 Jan 9

2. Hynninen J, Lavonius M, Oksa S, et al. Is perioperative visual estimation of intra-abdominal tumor spread reliable in ovarian cancer surgery after neoadjuvant chemotherapy? Gynecol Oncol. 2013; 128: 229-232, 2013

3. Hennessy BT, Coleman RL, Markman M (2009) Ovarian cancer. Lancet 374:1371–1382. https://doi.org/10.1016/S0140-6736(0961338-6)

4. Bhatt A, Glehen O (2016) the role of cytoreductive surgery and hyperthermic intraperitoneal chemotherapy (hipec) in ovarian cancer: a review. Indian J Surg Oncol 7(2):188–197. https://doi.org/10.1007/s13193-016-0501-9

5. Minig L, Guadalupe Patrono M, Alvarez Gallego R, et al (February 27th 2013). Surgical Treatment of Ovarian Cancer, Ovarian Cancer - A Clinical and Translational Update, Iván Díaz-Padilla, IntechOpen, DOI: 10.5772/53972. Available from: https://www.intechopen.com/books/ovarian-cancer-a-clinical-and-translational-update/surgical-treatment-of-ovarian-cancer

6. Buchsbaum HJ, Brady MF, Delgado G, et al. Surgical staging of carcinoma of the ovaries. Surg Gynecol Obstet. 1989;169(3):226–32.

7. Lim MC^1^, Song YJ, Seo SS, et al. Residual cancer stem cells after interval cytoreductive surgery following neoadjuvant chemotherapy could result in poor treatment outcomes for ovarian cancer. Onkologie. 2010;33(6):324–30. doi: 10.1159/000313823. Epub 2010 May 14.

8. Querleu D^1^, Planchamp F^1^, Chiva L^2^, et al. European Society of Gynaecological Oncology (ESGO) Guidelines for Ovarian Cancer Surgery. Int J Gynecol Cancer. 2017 Sep;27(7):1534–1542. doi: 10.1097/IGC.0000000000001041.

9. NCCN guidelines https://www.nccn.org/professionals/physician_gls/pdf/ovarian.pdf. Accessed 30th March, 2020

10. Jacquet P, Sugarbaker PH. Clinical research methodologies in diagnosis and staging of patients with peritoneal carcinomatosis. Cancer Treat Res. 1996; 82:359–374.

11. Bhatt A, Yonemura Y, Benzerdjeb N, et al. Pathological assessment of cytoreductive surgery specimens and its unexplored prognostic potential-a prospective multi-centric study. Eur J Surg Oncol. 2019 Dec;45(12):2398–2404

12. McCluggage WG, Judge MJ, Clarke BA, et al. International collaboration on cancer reporting. Data set for reporting of ovary, fallopian tube and primary peritoneal carcinoma: recommendations from the International Collaboration on Cancer Reporting (ICCR). 2015. Mod Pathol 28(8):1101–1122. https://doi.org/10.1038/modpathol.2015.77

13. Bhatt, A., Yonemura, Y., Mehta, S. et al. The Pathologic Peritoneal Cancer Index (PCI) Strongly Differs From the Surgical PCI in Peritoneal Metastases Arising From Various Primary Tumors. Ann Surg Oncol (2020). https://doi.org/10.1245/s10434-020-08234-x

14. Villeneuve L, Thivolet A, Bakrin N, et al. A new internet tool to report peritoneal malignancy extent. PeRitOneal MalIgnancy stage evaluation (PROMISE) application. Eur J Surg Oncol. 2016;42(6):877–82

15. Böhm S, Faruqi A, Said I, et al (2015) Chemotherapy response score: development and validation of a system to quantify histopathologic response to neoadjuvant chemotherapy in tubo-ovarian high-grade serous carcinoma. J Clin Oncol 33(22):2457–2463. https://doi.org/10.1200/JCO. 2014.60.5212

16. Sugarbaker PH (1995) Peritonectomy procedures. Ann Surg 221(1):29–42.

17. Sinukumar S, Rajan F, Mehta S, et al. A comparison of outcomes following total and selective peritonectomy performed at the time of interval cytoreductive surgery for advanced serous epithelial ovarian, fallopian tube and primary peritoneal cancer - A study by INDEPSO. Eur J Surg Oncol. 2019 Feb 23. pii: S0748-7983(19)30304-X. doi: 10.1016/j.ejso.2019.02.031.

18. Somashekhar, S., Ashwin, K., Yethadka, R., et al. (2019). Impact of extent of parietal peritonectomy on oncological outcome after cytoreductive surgery and HIPEC. Pleura and Peritoneum, 4(4), pp. -. Retrieved 25 Nov. 2019, from doi:10.1515/pp-2019-0015

19. Early Breast Cancer Trialists’ Collaborative Group (EBCTCG) Long-term outcomes for neoadjuvant versus adjuvant chemotherapy in early breast cancer: meta-analysis of individual patient data from ten randomised trials. Lancet Oncol. 2018;19(1):27–39. doi: 10.1016/S1470-2045(17)30777-5.

20. Karagkounis G^1^, Thai L^1^, Mace AG^1^, et al. Prognostic Implications of Pathological Response to Neoadjuvant Chemoradiation in Pathologic Stage III Rectal Cancer. Ann Surg. 2018 Feb 20. doi: 10.1097/SLA.0000000000002719. [Epub ahead of print]

21. Chi DS, Musa F, Dao F, et al (2012) An analysis of patients with bulky advanced stage ovarian, tubal, and peritoneal carcinoma treated with primary debulking surgery (PDS) during an identical time period as the randomized EORTC-NCIC trial of PDS vs neoadjuvant chemotherapy (NACT). Gynecol Oncol 124:10–14

22. Bhatt, A., Sinukumar, S., Rajan, F., et al. Impact of Radicality Versus Timing of Surgery in Patients with Advanced Ovarian Cancer (Stage III C) Undergoing CRS and HIPEC—a Retrospective Study by INDEPSO Indian J Surg Oncol (2019) 10(Suppl 1): 57. https://doi.org/10.1007/s13193-019-00875-z

23. P. Harter, J. Sehouli, D. Lorusso, et al. A randomized trial of lymphadenectomy in patients with advanced ovarian neoplasms, N. Engl. J. Med. 380 (2019) 822–832, https://doi.org/10.1056/NEJMoa1808424.

24. Amate P^1^, Huchon C, Dessapt AL, et al. Ovarian cancer: sites of recurrence. Int J Gynecol Cancer. 2013 Nov;23(9):1590–6. doi: 10.1097/IGC.0000000000000007.

25. Pereira A, Pérez-Medina T, Magrina JF, et al (2014) Correlation between the extent of intraperitoneal disease and nodal metastasis in node-positive ovarian cancer patients. Eur J Surg Oncol 40(8):917–924

26. Kammar P^1^, Bhatt A^2^, Anam J^1^, et al. Correlation Between Pelvic Peritoneal Disease and Nodal Metastasis in Advanced OvarianCancer:C an Intraoperative Findings Define the Need for Systematic Nodal Dissection? Indian J Surg Oncol. 2019 Feb;10 (Suppl 1):84–90. doi: 10.1007/s13193-019-00881-1. Epub 2019 Feb 7.

27. Di Giorgio A, Cardi M, Biacchi D, et al (2013) Depth of colorectal-wall invasion and lymph-node involvement as major outcome factors influencing surgical strategy in patients with advanced and recurrent ovarian cancer with diffuse peritoneal metastases. World J Surg Oncol 11:64

28. Dohan, A., Hoeffel, C., Soyer, P, et al. (2017), Evaluation of the peritoneal carcinomatosis index with CT and MRI. Br J Surg, 104: 1244–1249

